# Prediction of autism spectrum disorder diagnosis using nonlinear measures of language-related EEG at 6 and 12 months

**DOI:** 10.1101/2020.12.16.20248200

**Authors:** Fleming C. Peck, Laurel J. Gabard-Durnam, Carol L. Wilkinson, William Bosl, Helen Tager-Flusberg, Charles A. Nelson

## Abstract

Early identification of autism spectrum disorder (ASD) provides an opportunity for early intervention and improved outcomes. Use of electroencephalography (EEG) in infants has shown promise in predicting later ASD diagnoses and in identifying neural mechanisms underlying the disorder. Given the high co-morbidity with language impairment in ASD, we and others have speculated that infants who are later diagnosed with ASD have altered language learning, including phoneme discrimination. Phoneme learning occurs rapidly within the first postnatal year, so altered neural substrates either during or after the first year may serve as early, accurate indicators of later autism diagnosis. Using longitudinal EEG data collected during a passive phoneme task in infants with high familial risk for ASD, we compared predictive accuracy at 6-months (during phoneme learning) versus 12-months (after phoneme learning). Samples at both ages were matched in size and diagnoses (n=14 with later ASD; n= 40 without ASD). Using Pearson correlation feature selection and support vector machine with radial basis function classifier, 100% predictive diagnostic accuracy was observed at both ages. However, predictive features selected at the two ages differed and came from different scalp locations. We also report that performance across multiple machine learning algorithms was highly variable and declined when the 12-month sample size and behavioral heterogeneity was increased. These results demonstrate that speech processing EEG measures can facilitate earlier identification of ASD but emphasize the need for age-specific predictive models with large sample sizes in order to develop clinically relevant classification algorithms.

## Introduction

The past decade has witnessed a dramatic increase in the prevalence of autism spectrum disorder (ASD), a neurodevelopmental disorder that is characterized by social communication and repetitive and restrictive behaviors *(1)*. The Centers for Disease Control and Prevention estimate that one in 54 children has an ASD diagnosis *(2)*. Currently, ASD is diagnosed using behavioral measures, so a diagnosis cannot be made until toddlerhood or later when behavioral symptoms are reliably observable *(3)*. However, studies show that introducing intervention to children earlier in development leads to better intellectual and behavioral outcomes *(4, 5)*. Early identification of ASD is essential to early intervention. Therefore, a central focus for the field has been to develop identification tools using biological markers to facilitate earlier detection of ASD.

Neuroimaging measures provide strong candidate tools for early identification as they can be obtained from prenatal ages onwards. For example, several recent studies have used magnetic resonance imaging (MRI) data collected in infancy to predict ASD diagnoses *(6, 7)*. However, MRI has several notable drawbacks that make it less feasible as a general screening tool: it is expensive, infants must be scanned while asleep, and it is unsafe for certain populations (e.g., those with metal implants). Electroencephalography (EEG), on the other hand, may prove to be a more scalable tool, given its low cost, ease of acquisition in awake or sleeping infants, and the fact that there are no exclusion criteria for its use. Moreover, EEG is known to be sensitive to brain-related changes in ASD before behavioral symptoms are observable *(8-12)*. Initial efforts to predict ASD diagnoses using baseline (i.e. resting-state) EEG early in life have shown promise *(13-16)*. However, ASD diagnostic prediction using EEG recorded during tasks related to ASD symptoms has not yet been attempted, and it is possible that task-related EEG measures outperform prior baseline EEG-based classification.

In particular, language is frequently delayed or impaired in ASD *(17-21)*. Theories of delays in language acquisition associated with ASD postulate that atypical peak synaptic sensitivity *(22)* and cortical excitatory and inhibitory imbalance *(23)* disrupt the neural circuits that typically allow for language development. Therefore, focusing on the brain’s electrical activity during language processing may facilitate improved diagnostic prediction accuracy relative to baseline conditions. Notably, EEG has been used to measure differences in language processing in children with ASD who are older than 12 months *(24-26)*, providing evidence that EEG is sensitive to atypical neural processing of language stimuli in ASD.

In this context of predicting ASD diagnosis, focusing on language acquisition within the first 12 months may yield the earliest indicators. One such critical stage in language acquisition is learning to discriminate native from non-native speech sounds, which occurs during a sensitive period over the first year of life. That is, very young infants can discriminate between native and non-native phonemes better than adults, but they lose this ability over the first year of life as their phoneme perception is tuned to the language(s) that they experience in their daily lives (27). This perceptual narrowing of phoneme discrimination is necessary for speech perception and language learning *(28, 29)* and may be processed differently in infants with ASD *(30-32)*. Therefore, this study focused on the phoneme learning window over the first year of life as a potential source of early task-related EEG indicators of subsequent ASD diagnosis.

There were two overarching goals of the present study. First, we aimed to evaluate whether EEG data collected during a language phoneme task at either 6- or 12-months of age in infants with familial risk for ASD can accurately predict later ASD diagnosis. To do this, we utilized EEG data collected from high familial risk infant siblings as part of a prospective longitudinal study, where diagnosis of ASD was determined at 2-3 years of age. Specifically, power and nonlinear features of the EEG were tested as potential predictors of subsequent diagnosis. Though power analysis of EEG is most common, electrical activity recorded from the brain has complex dynamical properties that power analysis is not able to quantify. Moreover, these nonlinear features in adult EEG have accurately classified other clinical conditions, including depression *(33-35)*, schizophrenia *(36-39)*, and epilepsy *(40-42)*. Therefore, a combination of nonlinear measures that quantify signal complexity and regularity can describe different properties of the neural time series and may improve ASD prediction. Our lab has previously found that these measures computed from resting state EEG are useful in predicting ASD outcome *(15)*. Second, given that expected perceptual narrowing of phoneme discrimination occurs between 6 and 12 months of age, we next aimed to identify and compare the EEG features most predictive of diagnosis at each age and determine whether there are developmental differences in which features are most important during versus after the language phoneme learning period. To do this, 6- and 12-month datasets were matched in number as well as demographic and ASD outcome characteristics.

## Results

### Participants and data characteristics

Three sets of EEG data were evaluated in predictive models: 6-month sample, full 12-month sample, and a matching 12-month sample. The 6-month sample included a total of 54 high-risk infants, 40 of whom did not meet criteria for ASD (HR-NoASD) and 14 of whom were later diagnosed with ASD (HR-ASD). The full 12-month sample included a total of 67 high-risk infants (40 HR-noASD; 27 HR-ASD). In order to more directly compare 6- and 12-month data sets, we created a “matching” 12-month dataset with the same ratio of HR-noASD and HR-ASD infants (40 HR-NoASD vs 14 HR-ASD). These matched datasets included EEG from 39 infants who provided data at both 6- and 12-month times points (24 HR-noASD; 13 HR-ASD). 12-month EEG data from one additional HR-ASD infant was included in this matched dataset and was selected a priori based on matching demographic information of the one 6-month participant who developed ASD but did not participate at 12 months. Demographic characteristics of the infants in all three groups are provided in Table 4.

### Autism prediction at 6 months

We evaluated the ability of multiple feature selection methods and machine learning algorithms to predict ASD diagnosis. Three distinct feature selection methods were included: selection based on (1) the features most correlated with autism outcomes (Pearson correlation coefficient), (2) the features with most significant F ratio of mean square variances by group (F-test), and (3) the features selected using recursive feature elimination with cross validation based on a linear support vector machine (recursive feature elimination). We evaluated the prediction ability of four machine learning algorithms with different classification strategies: support vector machine (SVM) with radial basis function (discriminative classifier with a hyperboundary), *k*-nearest neighbors (voting method), linear discriminant analysis (linear combination of features), and Naïve Bayes (probabilistic classifier). Each of these methods uses a different procedure to select features or model outcomes, so we anticipated variability in the predictive capacity with different combinations.

Considering the sample size and imbalanced nature of the dataset, nested leave-one-out cross validation was used to evaluate the ability of power and nonlinear measures of language-related EEG signal to predict later ASD diagnosis. At each iteration of cross validation, one participant’s data was removed from the dataset, features were selected based on the properties of the remaining samples, and the machine learning model trained using those predictive features from the remaining samples to predict the outcome of the left-out sample. Thus, the feature selection procedures were repeated entirely at every iteration without dependence on the left-out sample. Models were limited to 20 features to prevent over-fitting to the available data samples given the sample size. Table 1 shows the classification results for each of the feature selection methods and classifiers detailed above. Using 20 features selected by the Pearson correlation ranking method, the SVM classifier achieved 100% diagnostic prediction accuracy. Given the imbalanced nature of the sample, permutation testing was used to assess the significance of observed prediction accuracy. A null distribution of predictive accuracy was generated by repeating the following procedure over 1000 iterations: diagnostic labels were shuffled and cross validation analysis using features selected with true labels was performed. In the context of this null distribution, predictive accuracy using true labels was significantly better than chance (z-score 7.35, p <0.0001). Classification performance varied across combinations of the three feature selection, and the four machine learning methods, with an overall accuracy range of 20.4% to 100%. SVM performance was the most variable of all machine learning algorithms considered when used with different feature selection methods.

**Table 1:**
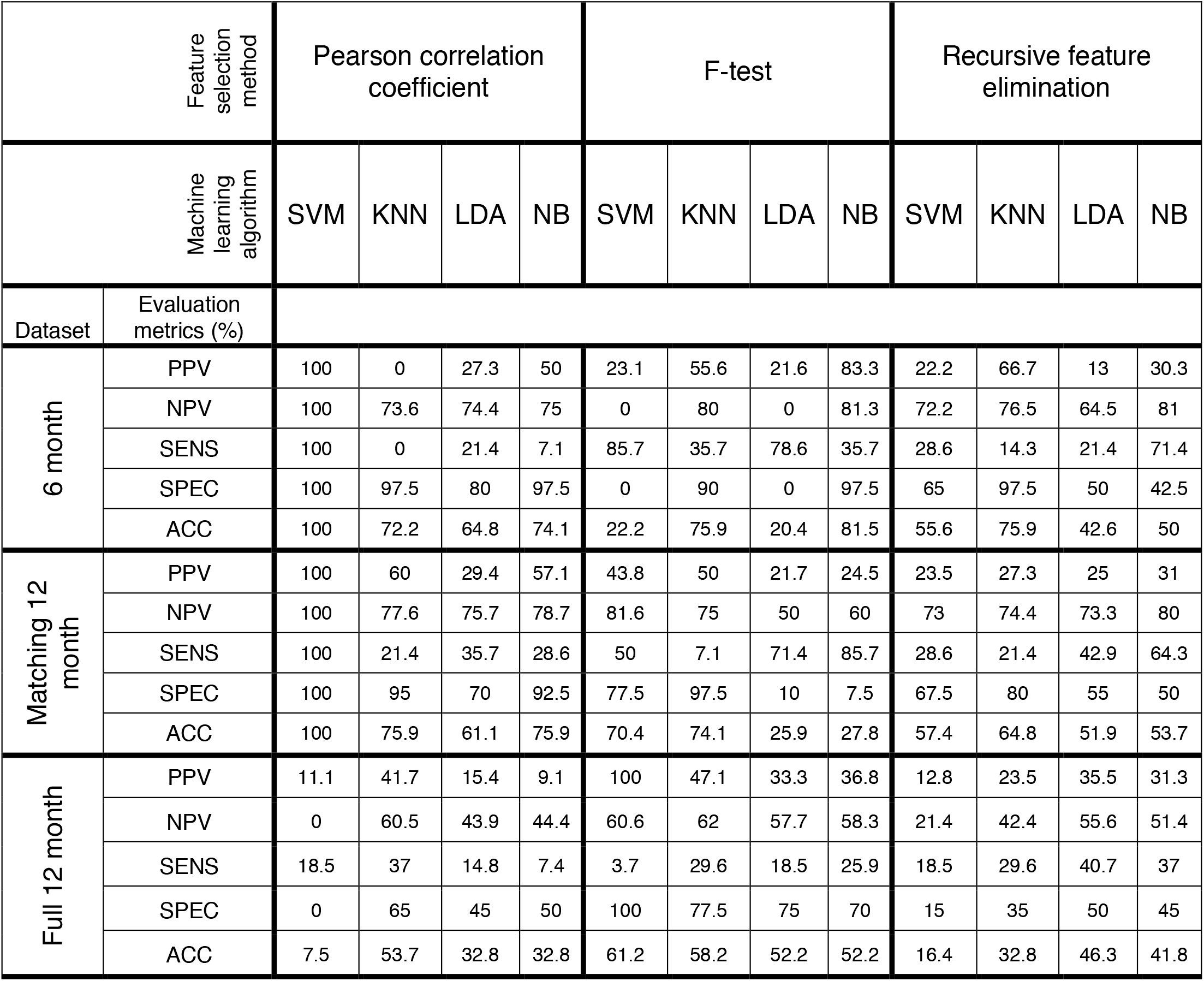
Evaluation metrics for machine learning classification results reported in percentages. All three datasets are reported: 6-month dataset (n = 54; 74.1% HR-noASD), matching 12-month dataset (n = 54; 74.1% HR-noASD), and full 12-month dataset (n = 67; 59.7% HR-noASD). Machine learning classification was evaluated in terms of positive predictive value (PPV), negative predictive value (NPV), sensitivity (SENS), specificity (SPEC), and accuracy (ACC). Evaluation metrics are included for four algorithms: nonlinear support vector machine (SVM), K-nearest neighbors with 7 neighbors (KNN), linear discriminant analysis (LDA), and Naïve Bayes (NB). 20 features were selected by the noted selection methods before nested leave-one-out cross validation using each of the listed

### Autism prediction at 12 months: matched sample

In order to compare ASD prediction accuracy at 6 months (during the phoneme learning period) and 12 months (the end of the phoneme learning period), and to compare the nature of the features selected for diagnosis prediction at these two ages, we next analyzed our “matched” 12-month dataset with the same total number of infants and the same proportion of HR-noASD and HR-ASD infants as the 6-month dataset. We evaluated the same feature selection methods and classifiers as in the previous analysis, and the nested cross validation results are reported in Table 1. 100% classification accuracy was achieved using feature selection based on correlation to ASD outcomes and an SVM classifier—the same feature selection method and machine learning algorithm that achieved the most successful prediction accuracy at 6-months. Evaluation metrics based on features selected by the Pearson correlation coefficient were higher than the other two feature selection methods for all evaluation metrics except sensitivity across all machine learning algorithms. Overall, classification accuracies ranged from 25.9 to 100%. Using permutation testing described above, the 100% classification accuracy was significantly better than chance (z-score 7.5, p<0.0001). As with the 6-month prediction results, sensitivity rates were generally lower across models than specificity rates.

### Autism prediction at 12 months: full sample

To test the prediction consistency across samples, we evaluated the prediction accuracy of models in the full 12-month dataset, which had 13 additional HR-ASD participants (40 HR-noASD and 27 HR-ASD) who only participated at 12 months of age, almost doubling the HR-ASD sample. The classification results are presented in Table 1. There was a considerable decrease in almost all evaluation metrics and especially overall prediction accuracy from the previous two classification reports. Only one of the 12 classification schemes presented in Table 1—F-test feature selection with SVM algorithm—resulted in accuracy marginally above chance (61.2%) at the severe expense of sensitivity (3.7%), the measure evaluating the percentage of infants with ASD who were predicted correctly.

Sensitivity was the lowest evaluation metric for the full 12-month dataset for nearly all of the classification schemes that we evaluated. Like the 6-month and matched 12-month analyses, sensitivity was almost always lower than specificity. However, the poor prediction across models resulted from poor performance across all evaluation metrics, rather than a consistent under- or over-prediction of ASD diagnosis.

To investigate possible reasons for the differences in classification accuracy between the matched and full 12-month analyses, we assessed whether there were differences in behavioral measures of the HR-ASD participants. We evaluated five different behavioral assessments and used a *t*-test to compare performance of HR-ASD infants included in the matched data set versus HR-ASD only included in the 12-month full dataset (Table 2). We found that the 12-month participants who participated at the 6-month timepoint had significantly lower severity scores on the Autism Diagnostic Observation Schedule (ADOS) at 36 months compared to the non-matching 12-month infants who only contributed EEG data at 12 months of age. In addition, there were no significant differences between samples in terms of EEG length or quality using HAPPE metrics (all *p* > 0.1).

**Table 2:** Behavioral assessments of 12-month HR-ASD group by participation timepoints. P-value of t-test comparing scores of each behavioral assessment between the matching and nonmatching 12-month HR-ASD infants (significant p-value is emboldened). ADOS: Autism Diagnostic Observation Schedule. MSEL: Mullen Scales of Early Learning.

|  | Full HR-ASD | Matching HR-ASD | Nonmatching HR-ASD | p-value |
| --- | --- | --- | --- | --- |
| Behavioral assessments<br>(mean ± SD (n)) | | | | |
| ADOS |  |  |  |  |
| 24mo Severity Score | 4.81 ± 2.5<br>(25) | 4.21 ± 2.29<br>(14) | 5.81 ± 2.79<br>(11) | 0.15 |
| 36mo Severity Score | 4.9 ± 2.19<br>(23) | 3.85 ± 1.68<br>(13) | 6.3 ± 2.93<br>(10) | **0.006** |
| MSEL | n = 23 | n = 12 | n = 11 |  |
| Composite Scaled Score | 97.5 ± 15.6 | 101.5 ± 13.9 | 93.3 ± 16.9 | 0.21 |
| Verbal Developmental Quotient | 88.9 ± 21.8 | 92.7 ± 21.5 | 84.7 ± 22.3 | 0.39 |
| Non-verbal Developmental Quotient | 114.9 ± 12 | 118.7 ± 10.4 | 110.7 ± 14.4 | 0.14 |

### Predictive features at 6 and 12 months

Finally, the nature and spatial distribution of features selected in the successful 6-month and 12-month predictive models were extracted in order to compare EEG features most predictive of ASD diagnosis either during (6 months of age) or after (12 months of age) perceptual narrowing of phoneme discrimination. Importantly, the same feature selection method and machine learning algorithm (Pearson correlation coefficient feature selection and an SVM classifier) achieved 100% predictive accuracy for both the 6-month and matched 12-month datasets, allowing for direct comparison of the features selected at each iteration of nested cross validation across the two ages. Given poor prediction of full 12-month sample, we do not report the features for this sample.

Figure 1 shows the selection rates of features by channel, measure, and frequency. At 6 months, features were selected largely from central and left of the midline locations (Fig 1A), and power was the most frequently selected measure (Fig 1B). Five of the 12 measures (power, approximate entropy, Hurst exponent, Lempel-Ziv complexity, and permutation entropy were consistently selected across iterations. Within iterations, power was the most frequently selected feature and made up the majority of the 20 selected features. While measures related to all frequency bands were consistently chosen across iterations, the average count per iteration of features related to the 15.6-31.2 Hz range (largely canonical Beta frequencies) was nearly double any other frequency range (Fig 1C). At 12 months, selected channels changed most in the left hemisphere, with increased left-lateralization (i.e. shifts away from midline) and representation from especially dense frontal and temporo-parietal regions (Fig 1D). Seven of the 12 measures were selected in at least 80% of the iterations, and half of the selected measures in each iteration were power or Lyapunov exponent computed at different wavebands and channels (Fig 1E).

**Fig. 1.**
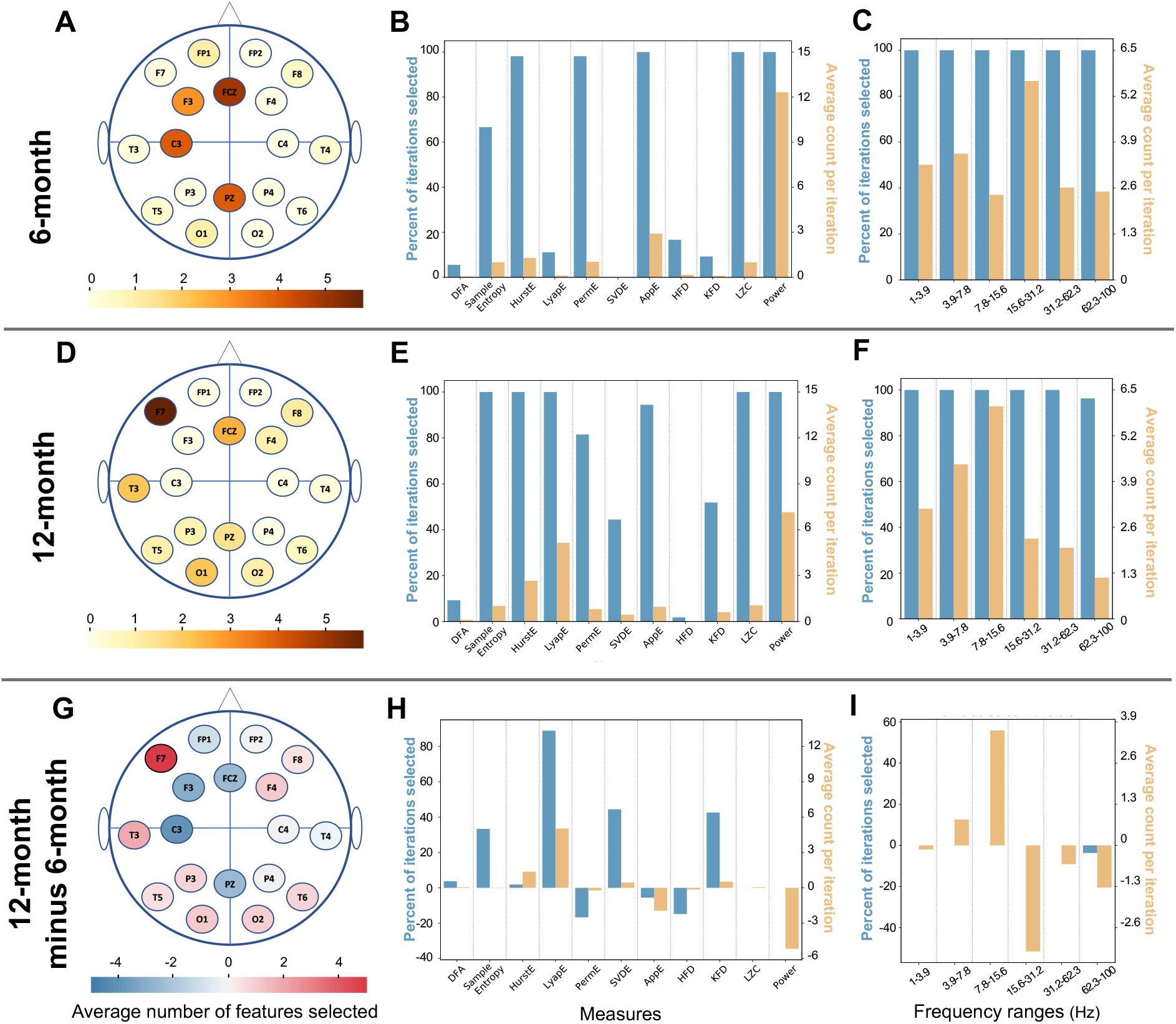
Information about features most correlated with autism diagnostic outcome for nearly overlapping 6- and 12-month analyses (n = 54). The bottom row visualizes the values for the 12-month dataset (middle row) minus the 6-month dataset (top row). (**A, D, G**) Average number of features selected from each channel. Color indicates number of features selected from a given channel. (**B, E, H**) Average count of each EEG measure across iterations (orange) and percentage of iterations that each measure was selected at least once (blue). (**C, F, I**) Average count of each wavelet across iterations (orange) and percentage of iterations that each wavelet was selected at least once (blue).

Additionally, while almost all frequency bands were selected at each iteration, signal filtered to frequency ranges below 16 Hz were more frequently selected at age 12 months (Fig 1F). We next compared the mean value of each of the 20 most frequently selected features between outcome groups (HR-ASD and HR-noASD; Table 3). After correcting for multiple comparisons, only approximate entropy computed from the F3 electrode in the delta range (1-4Hz) was significantly different between the two groups at 6-months, with HR-ASD demonstrating higher entropy than the HR-noASD infants (HR-ASD: mean 0.815± 0.08; HR-noASD: mean 0.76 ± 0.037; p <0.0025). In contrast, at 12 months of age, the mean values measured for seven of the 20 features most commonly chosen across iterations were significantly different between HR-ASD and HR-noASD groups such that HR-ASD infants had consistently higher values for each of these features than HR-noASD infants. Significant features across model iterations were also those that were most often chosen within a given iteration’s set of 20 features (Fig 1E): Lyapunov exponent, Hurst Exponent, sample entropy, and power. Lempel-Ziv complexity was the only measure selected in all iterations that was not significantly different between groups after Bonferroni correction.

**Table 3.**
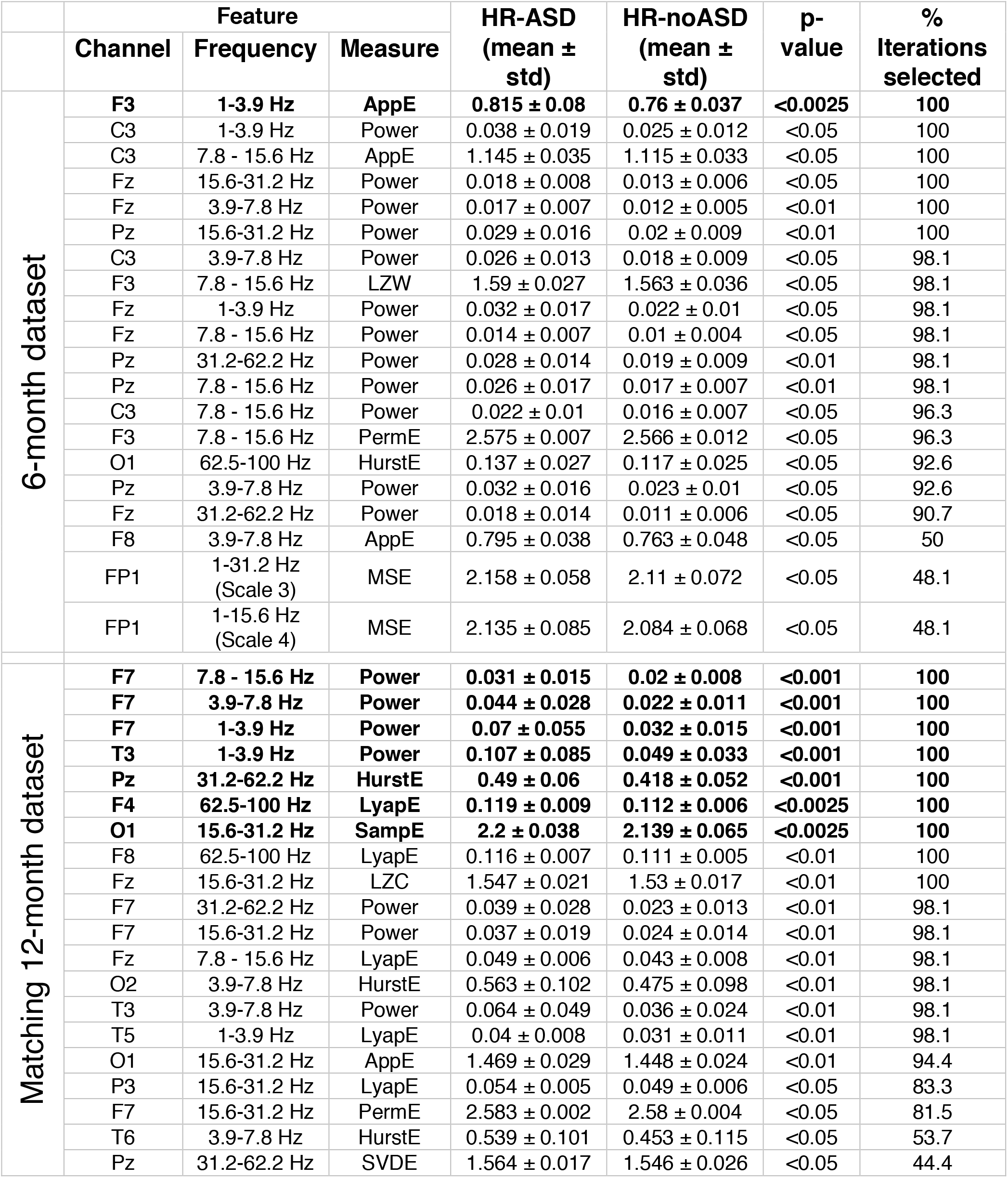
Descriptions of the 20 most-frequently selected features using the Pearson Correlation Coefficient features selection method. Scale in the frequency band feature category refers to the level of coarse-graining procedure (further described in Methods). Significance evaluated with paired sample t-test corrected for 20 comparisons. Parameters that survived Bonferroni correction are in bold

| | Feature | | | HR-ASD<br>(mean ± std) | HR-noASD<br>(mean ± std) | p-value | %<br>Iterations<br>selected |
| --- | --- | --- | --- | --- | --- | --- | --- |
|  | Channel | Frequency | Measure |  |  |  |  |
| 6-month dataset | **F3** | **1-3.9 Hz** | **AppE** | **0.815 ± 0.08** | **0.76 ± 0.037** | **<0.0025** | **100** |
| | C3 | 1-3.9 Hz | Power | 0.038 ± 0.019 | 0.025 ± 0.012 | <0.05 | 100 |
| | C3 | 7.8 - 15.6 Hz | AppE | 1.145 ± 0.035 | 1.115 ± 0.033 | <0.05 | 100 |
| | Fz | 15.6-31.2 Hz | Power | 0.018 ± 0.008 | 0.013 ± 0.006 | <0.05 | 100 |
| | Fz | 3.9-7.8 Hz | Power | 0.017 ± 0.007 | 0.012 ± 0.005 | <0.01 | 100 |
| | Pz | 15.6-31.2 Hz | Power | 0.029 ± 0.016 | 0.02 ± 0.009 | <0.01 | 100 |
| | C3 | 3.9-7.8 Hz | Power | 0.026 ± 0.013 | 0.018 ± 0.009 | <0.05 | 98.1 |
| | F3 | 7.8 - 15.6 Hz | LZW | 1.59 ± 0.027 | 1.563 ± 0.036 | <0.05 | 98.1 |
| | Fz | 1-3.9 Hz | Power | 0.032 ± 0.017 | 0.022 ± 0.01 | <0.05 | 98.1 |
| | Fz | 7.8 - 15.6 Hz | Power | 0.014 ± 0.007 | 0.01 ± 0.004 | <0.05 | 98.1 |
| | Pz | 31.2-62.2 Hz | Power | 0.028 ± 0.014 | 0.019 ± 0.009 | <0.01 | 98.1 |
| | Pz | 7.8 - 15.6 Hz | Power | 0.026 ± 0.017 | 0.017 ± 0.007 | <0.01 | 98.1 |
| | C3 | 7.8 - 15.6 Hz | Power | 0.022 ± 0.01 | 0.016 ± 0.007 | <0.05 | 96.3 |
| | F3 | 7.8 - 15.6 Hz | PermE | 2.575 ± 0.007 | 2.566 ± 0.012 | <0.05 | 96.3 |
| | O1 | 62.5-100 Hz | HurstE | 0.137 ± 0.027 | 0.117 ± 0.025 | <0.05 | 92.6 |
| | Pz | 3.9-7.8 Hz | Power | 0.032 ± 0.016 | 0.023 ± 0.01 | <0.05 | 92.6 |
| | Fz | 31.2-62.2 Hz | Power | 0.018 ± 0.014 | 0.011 ± 0.006 | <0.05 | 90.7 |
| | F8 | 3.9-7.8 Hz | AppE | 0.795 ± 0.038 | 0.763 ± 0.048 | <0.05 | 50 |
| | FP1 | 1-31.2 Hz<br>(Scale 3) | MSE | 2.158 ± 0.058 | 2.11 ± 0.072 | <0.05 | 48.1 |
| | FP1 | 1-15.6 Hz<br>(Scale 4) | MSE | 2.135 ± 0.085 | 2.084 ± 0.068 | <0.05 | 48.1 |
| Matching 12-month dataset | **F7** | **7.8 - 15.6 Hz** | **Power** | **0.031 ± 0.015** | **0.02 ± 0.008** | **<0.001** | **100** |
|  | **F7** | **3.9-7.8 Hz** | **Power** | **0.044 ± 0.028** | **0.022 ± 0.011** | **<0.001** | **100** |
|  | **F7** | **1-3.9 Hz** | **Power** | **0.07 ± 0.055** | **0.032 ± 0.015** | **<0.001** | **100** |
|  | **T3** | **1-3.9 Hz** | **Power** | **0.107 ± 0.085** | **0.049 ± 0.033** | **<0.001** | **100** |
|  | **Pz** | **31.2-62.2 Hz** | **HurstE** | **0.49 ± 0.06** | **0.418 ± 0.052** | **<0.001** | **100** |
|  | **F4** | **62.5-100 Hz** | **LyapE** | **0.119 ± 0.009** | **0.112 ± 0.006** | **<0.0025** | **100** |
|  | **O1** | **15.6-31.2 Hz** | **SampE** | **2.2 ± 0.038** | **2.139 ± 0.065** | **<0.0025** | **100** |
| | F8 | 62.5-100 Hz | LyapE | 0.116 ± 0.007 | 0.111 ± 0.005 | <0.01 | 100 |
| | Fz | 15.6-31.2 Hz | LZC | 1.547 ± 0.021 | 1.53 ± 0.017 | <0.01 | 100 |
| | F7 | 31.2-62.2 Hz | Power | 0.039 ± 0.028 | 0.023 ± 0.013 | <0.01 | 98.1 |
| | F7 | 15.6-31.2 Hz | Power | 0.037 ± 0.019 | 0.024 ± 0.014 | <0.01 | 98.1 |
| | Fz | 7.8 - 15.6 Hz | LyapE | 0.049 ± 0.006 | 0.043 ± 0.008 | <0.01 | 98.1 |
| | O2 | 3.9-7.8 Hz | HurstE | 0.563 ± 0.102 | 0.475 ± 0.098 | <0.01 | 98.1 |
| | T3 | 3.9-7.8 Hz | Power | 0.064 ± 0.049 | 0.036 ± 0.024 | <0.01 | 98.1 |
| | T5 | 1-3.9 Hz | LyapE | 0.04 ± 0.008 | 0.031 ± 0.011 | <0.01 | 98.1 |
| | O1 | 15.6-31.2 Hz | AppE | 1.469 ± 0.029 | 1.448 ± 0.024 | <0.01 | 94.4 |
| | P3 | 15.6-31.2 Hz | LyapE | 0.054 ± 0.005 | 0.049 ± 0.006 | <0.05 | 83.3 |
| | F7 | 15.6-31.2 Hz | PermE | 2.583 ± 0.002 | 2.58 ± 0.004 | <0.05 | 81.5 |
| | T6 | 3.9-7.8 Hz | HurstE | 0.539 ± 0.101 | 0.453 ± 0.115 | <0.05 | 53.7 |
| | Pz | 31.2-62.2 Hz | SVDE | 1.564 ± 0.017 | 1.546 ± 0.026 | <0.05 | 44.4 |

## Discussion

In this study we evaluated multiple classification schemes to predict ASD diagnosis in high-familial risk infants using EEG data from infancy. Specifically, EEG signals collected during a language phoneme task were evaluated at either 6- or 12-months of age, timepoints that span a critical early language-learning period in development. 100% diagnostic classification accuracy was achieved for both 6- and matched 12-month data sets using the Pearson correlation coefficient feature selection method with the SVM classifier. That is, the same prediction rate was achieved from phoneme-related EEG regardless of whether infants were within the critical period of language phoneme learning (6 months) or after (12 months). However, the features selected to achieve the 100% prediction rate were quite different between the two ages, both in feature type and spatial distribution. Importantly, across classification schemes at both ages, model performance was highly variable and notably reduced when the sample demographics and size changed. Although we used robust statistical methods to limit overfitting in small samples, these constrained models may be unable to fit the variability present in ASD. The implications of these results in the search for early neuroimaging biomarkers of ASD diagnosis are discussed below.

### Developmental shift in predictive features

The longitudinal nature of the study provides the opportunity to assess whether predictive EEG features change across the course of a critical early period of language-learning within the first year of life. Several differences were identified. First, at 6 months power dominated as the most common measure selected, whereas at 12 months additional nonlinear measures were more consistently selected (Fig 1H). This trend is consistent with previous longitudinal studies that have found power differences between high- and low-risk infants that emerge before 6 months of age but often dissipate by 12 months of age, and can differentiate later ASD diagnostic outcomes *(8, 10, 13, 43)*. We also observed differences between developmental time points topographically, with a noticeable shift leftwards from central-midline locations, including increased involvement of the temporal lobe and lateral frontal cortex at 12 months (Fig 1G). This shift may indicate that infants who go on to have an ASD diagnosis show atypical language processing in the network of left-lateralized regions involved in healthy language perception by 12 months of age *(44-48)*. Moreover, these early differences are robust enough to indicate subsequent diagnostic outcome. Our EEG-related findings corroborate MRI findings of atypical activation related to passive auditory stimuli in adults and children with autism over similar cortical areas *(44-51)*.

Interestingly, the way features contributed to prediction shifted across development as well. At 12 months, more individual features were significantly different between ASD outcome groups (7 features) compared to 6 months (just 1 feature). One possible explanation is different modes of classification are utilized at the two ages, with a combination of feature values taken together at 6 months and independent contributions of individual features at 12 months. For example, power differences between ASD outcome groups were larger and more often statistically significant at 12 months, though these features were selected more frequently at 6 months (Table 2). At 12 months, power computed from the F7 channel was particularly predictive: all five frequency ranges from which power was computed were in the 20 most commonly selected features, and power was significantly different between HR-ASD and HR-noASD groups for the three frequency ranges below 32 Hz. Previous work suggests a positive correlation between language and cognitive outcomes and low gamma power during both resting-state *(45, 52-55)* and auditory-task EEG *(44, 56, 57)*. However, the reduced selection of power features and features derived from high frequency signal at 12 months compared to 6 months (Figure 1) suggest that nonlinear measures of low frequency signal better define binary ASD diagnostic outcomes based on language task EEG in the first year of life.

### Variable classification performance

The nested leave-one-out cross validation results varied greatly between the different feature selection method and machine learning algorithm combinations. The variability in performance may be attributed to the small sample size of the datasets, which risk overfitting or underfitting to the training data despite our efforts to minimize the dimensionality of the data before classification. Similarly-sized studies using MRI data have previously demonstrated sensitivity lower than 90% with near-perfect specificity *(6, 7)* of ASD diagnosis prediction in high-familial risk infants in contrast to 100% sensitivity and specificity of the best performing models based on the 6-month and size-marched 12-month datasets of this study. Previously reported MRI studies only presented results from a single classification scheme, so it is unclear whether predictive accuracy was more or less consistent across classification schemes for MRI data.

The decrease in accuracy from 100% with the matched 12-month dataset to 7% with the full 12-month dataset using a SVM classifier suggests an inability to effectively separate the two diagnostic outcome classes after the HR-ASD group was expanded. We found behavioral phenotypes of the HR-ASD infants added to complete the full 12-month analysis to be more variable and severe than those who participated at both 6 and 12 months. It is possible that enrollment bias influenced the sample characteristics in that high-risk families enrolling at a later age may have had increased concerns about ASD related to observed symptoms. We postulate that the inclusion of more samples resulted in increased feature overlap across outcome classes, preventing accurate hyperboundary separation. Our 12-month results also suggest that more complex modeling will be required at this age to appropriately account for the full range of heterogeneity in ASD at the brain and behavioral levels. It is also possible that classification with these simpler machine learning approaches might do well at 6 months even with a bigger sample size if EEG measures tend to be less variable and better stratified between the ASD diagnostic outcomes at this younger age.

### The effect of heterogeneity and sample size on prediction

Autism is a heterogenous disorder—its defining categories are broad and encompass a spectrum of symptom severity. The ideal model for autism diagnosis prediction will be robust to the degree of variability or heterogeneity of the disorder. Our SVM-based classification results at 6 months and with matched samples at 12 months are highly accurate, but the decline in classification accuracy with increased sample size and important shifts in sample phenotypes (including ADOS severity) highlights factors that must be considered by the greater autism research community for future diagnosis prediction efforts. This within-study classification distinction serves as a case study of poor generalizability to a larger sample.

The HR-ASD infants who were included in both matched and full 12-month classification analyses had significantly different ADOS severity scores at 36 months than the HR-ASD infants who joined the study at the 12-month timepoint. On all five behavioral measures evaluated in Table 3, the 12-month-only HR-ASD group had higher ADOS scores and lower MSEL scores (corresponding to overall lower indexes of development). We hypothesize that the inclusion of a more severe and variable dataset reduced the accuracy of classification since we were predicting ASD as a binary diagnosis. Using resting-state EEG from the same infant-sibling dataset in this analysis, we have previously observed a similar lower accuracy in a 12-month sample, although a similar size sample at 9 months had high accuracy. As discussed in our previous paper, this may be a real neurodevelopmental trend *(14, 15)*. This finding is in line with previous meta-analyses within the brain-disorder field that have found decreasing accuracies reported as sample sizes—and, importantly, heterogeneity within the sample—increase *(58-60)*. However, this may also be a developmental neurophysiology effect that reflects the cross-over of neurodevelopmental trajectories of infants who do and do not go on to develop ASD. More sample data are needed in order to more completely represent the potentially subtle or variable brain activity differences that arise in ASD. Increased sample sizes also permit the use of more complex models that may more appropriately account for the variability and complex associations between brain activity and diagnosis.

### Limitations and future directions

In addition to the discussed challenges of sample size and heterogeneity, we acknowledge several limitations of the current study. First, this analysis focused on infants with familial risk of autism, a finding that may generalize to other ASD-risk groups or to the general infant population. Second, the specificity of our findings to ASD (versus other comorbid conditions such as language delay) is not known. Further testing across clinical populations (eg. Global developmental delay without ASD or isolated Language Delay) would be helpful in understanding the specificity of our findings and whether EEG could also be used to predict comorbidities with significant impact on functional outcomes. This study determines ASD outcome at age 3, which is appropriate for assessing ASD but not for many other developmental conditions that emerge across early childhood. Therefore, questions about comorbidity call for the extension of longitudinal studies across infancy to track participants beyond 3 years in order to capture a more complete and confident clinical description of participants. Additionally, our sample was not diverse ethnically, racially, or in income level. Predictive analyses require not only large sample sizes but must include infants from diverse populations in order to improve clinical applicability for the general population. These results suggest that collaboration across samples is critical to moving forward in developing early predictive models. Future studies of early predictive markers of ASD and other neurodevelopmental disorders need to be acutely aware of participant age, given the dramatic developmental changes in predictive feature profiles over the 6-month age window in the study. Moreover, given the variability of behavioral measures within the ASD outcome group, future studies should consider distinguishing different subpopulations or subprofiles of ASD grouped by biological presentations or phenotype profiles at the behavioral level as opposed to only binary diagnosis.

## Materials and Methods

### Study design and participant

Participants in this study were recruited to Boston Children’s Hospital/Harvard Medical School and Boston University to participate in a longitudinal study of brain and behavioral development of infant siblings of children with ASD. Institutional review board approval was obtained from Boston University and Boston Children’s Hospital (# X06-08-0374) prior to the start of the study. All infants had a gestational age of at least 36 weeks and no history of seizures, prenatal drug exposure, hearing impairment, or known genetic mutation involved in neurodevelopment. The infants were designated high risk (HR) for ASD based on the confirmed ASD clinical diagnosis of an older sibling. A total of 104 HR infants were enrolled in the longitudinal study. ASD outcome was determined using the Autism Diagnostic Observation Schedule (ADOS) in conjunction with a clinical best estimate. For infants meeting criteria on the ADOS or coming within 3 points of cutoffs, near the clinical diagnosis cutoff, a Licensed Clinical Psychologist reviewed scores and video recordings and provided a best estimate clinical judgment of three categories: typically developing, ASD, or non-spectrum disorder, such as anxiety or language delay. Of the 54 HR infants who met the inclusion criteria at 6 months, 14 infants developed ASD (HR-ASD) and 40 infants did not (HR-noASD). Data from 67 HR infants were included at 12 months (27 HR-ASD and 40 HR-noASD). 13 HR-ASD and 24 HR-noASD infants were included at both 6- and 12-month timepoints. In order to assess longitudinal changes in ASD prediction at different ages, a “matching” 12-month dataset was curated by including data from the HR-ASD and HR-noASD infants who participated at 6 and 12 months. One HR-ASD participant from the 6-month cohort did not contribute data at 12 months, so a corresponding match was identified based on demographic similarity to replace that case. All 40 HR-ASD 12-month infant samples were included in the “matching” 12-month dataset, resulting in the same sample size as that of the 6-month dataset. Demographic and EEG data quality information of each outcome groups contributing data is presented in Table 4. Fisher’s exact test was used to evaluate significant differences of demographic information between groups. The 12-month high-risk group with ASD had significantly lower mean maternal education than the high-risk group without ASD *(p* = 0.004). No other demographic measures were significantly different between groups.

**TABLE 4.** Sample demographics of 6- and 12-month-old participants.

|  | 6-month dataset |  | 12-month dataset |  |  |
| --- | --- | --- | --- | --- | --- |
|  | **HR-ASD**<br>**n = 14** | **HR-noASD**<br>**n = 40** | **HR-ASD**<br>**n = 27** | **Matching**<br>**HR-ASD**<br>**n = 14** | **HR-noASD**<br>**n = 40** |
| Sex | 8 M, 6 F | 19 M, 21 F | 17 M, 10 F | 8 M, 6 F | 18 M, 22 F |
| Child ethnicity (%) |  |  |  |  |  |
| Caucasian | 92.9 | 97.5 | 77.8 | 92.9 | 95 |
| Hispanic/Latinx | 21.4 | 2.5 | 14.8 | 21.4 | 7.5 |
| Asian American | 0 | 0 | 3.7 | 0 | 0 |
| African American | 0 | 0 | 3.7 | 0 | 0 |
| Multirace | 7.1 | 2.5 | 14.8 | 7.1 | 7.1 |
| Mean household income (\$1000s) | 65-75 | 65-75 | 65-75 | 65-75 | 65-75 |
| Mean maternal education (%) |  |  |  |  |  |
| < 4-year college | 28.6 | 27.5 | 37 | 28.6 | 25 |
| = 4-year college | 35.7 | 15 | 37 | 42.9 | 12.5 |
| > 4-year college | 35.7 | 57.5 | 25.9 | 28.5 | 62.5 |
| EEG HAPPE metrics (mean [SD]) |  |  |  |  |  |
| Length of raw EEG (seconds) | 701.3 [212.9] | 693.6 [220.7] | 763.7 [213] | 735.8 [225.2] | 764.8 [209.5] |
| Good channels (%) | 93.1 [4.1] | 94.2 [4.7] | 92 [4.8] | 91.6 [4.7] | 92.8 [5.5] |
| Rejected components (%) | 47 [13] | 44.1 [13] | 43.8 [10] | 42.6 [8.4] | 43.4 [12.2] |
| EEG variance retained (%) | 61.6 [13.3] | 64.9 [17.4] | 66.5 [12.6] | 70.1 [10.5] | 69.6 [16] |
| Mean retained artifact probability | 0.16 [0.05] | 0.17 [0.03] | 0.15 [0.05] | 0.16 [0.05] | 0.15 [0.04] |
| Median retained artifact probability | 0.13 [0.06] | 0.15 [0.06] | 0.1 [0.07] | 0.11 [0.07] | 0.1 [0.06] |

### Behavioral assessments

The Mullen Scales for Early Learning (MSEL) was administered at each data collection visit of the longitudinal study, including 6 and 12 months. The MSEL provides an index of ability in domains including language, cognition, and motor development. Additionally, the Autism Diagnostic Observation Schedule (ADOS) was administered at 24- and 36-month visits. The MSEL composite score and verbal and non-verbal developmental quotients at 6 and 12 months, and ADOS calibrated severity scores at 24 and 36 months were used to evaluate developmental differences within the HR-ASD group.

### EEG paradigm

The phoneme speech task consisted of a stream of consonant-vowel stimuli that were presented using a double-oddball paradigm. Three phonemes were included and presented based on a probability of selection: the English standard “/da/” presented 80% of the time, the native deviant “/ta/” presented 10% of the time, and the non-native deviant “da/” presented 10% of the time. A maximum of 600 stimuli were presented with a variable interstimulus period of minimum 700ms. The language task lasted on average 15 minutes. For this study, only stretches of consecutive standard native phonemes (English ‘da’) presentations were included to ensure consistent stimuli ordering across individuals and so that the features were calculated over a single stimulus condition (no oddball stimuli were analyzed in the study).

### EEG data acquisition

EEG data were acquired in a dimly lit, sound-attenuated, electrically-shielded room. A research assistant was present in the room to ensure that the infant remained calm and still during the language paradigm by blowing bubbles or presenting toys if the infant became distracted or fussy. Assistants did not engage in social interaction with the infant during task completion. EEG data were collected with either a 64-channel Geodesic Sensor Nets or a 128-channel Hydrocel Geodesic Senso Net (Electrical Geodesics, Inc (EGI), Eugene, OR, USA), using a 0.1 Hz high-pass analog filter and online rereferencing to the vertex (channel Cz) through NetStation software (EGI, Eugene, OR, USA). Impedances were kept below 100KΩ in accordance with the connected DC-coupled amplifier (Net Amps 200 or Net Amps 300, Electrical Geodesics, Inc.). Data were sampled at either 250 or 500 Hz.

The EEG data were exported from NetStation to MATLAB format (R2017A). All files were batch processed using the Batch Electroencephalography Automated Processing Platform (BEAPP) that allows for preprocessing, artifact removal, and data quality assessment by the Harvard Automated Processing Pipeline for EEG (HAPPE) with the same empirical criteria for artifact removal applied in the same way across all files in the batch *(61)*. HAPPE is a preprocessing pipeline optimized for developmental EEG data, which has been shown to reject more artifacts and preserve more EEG signal during processing *(62)*. A spatially distributed subset of channels providing whole-head coverage were processed through HAPPE in order to optimize artifact rejection performance given the duration and sampling rates of the EEG data. This consisted of the 10-20 montage channel equivalents for each net type and channels clustered over the bilateral temporal-parietal cortex (64-channel net—2, 3, 6, 8, 9, 11, 12, 13, 15, 16, 17, 21, 24, 25, 27, 28, 34, 37, 40, 46, 49, 50, 52, 53, 54, 57, 58, 61, 62; 128-channel net—3, 4, 9, 11, 13, 19, 20, 22, 23, 24, 27, 28, 33, 36, 40, 41, 45, 46, 47, 52, 58, 62, 70, 75, 83, 92, 96, 98, 102, 103, 104, 108, 109, 112, 117, 118, 122, 123, 124). See Figure 2 for net layouts.

**Fig. 2.**
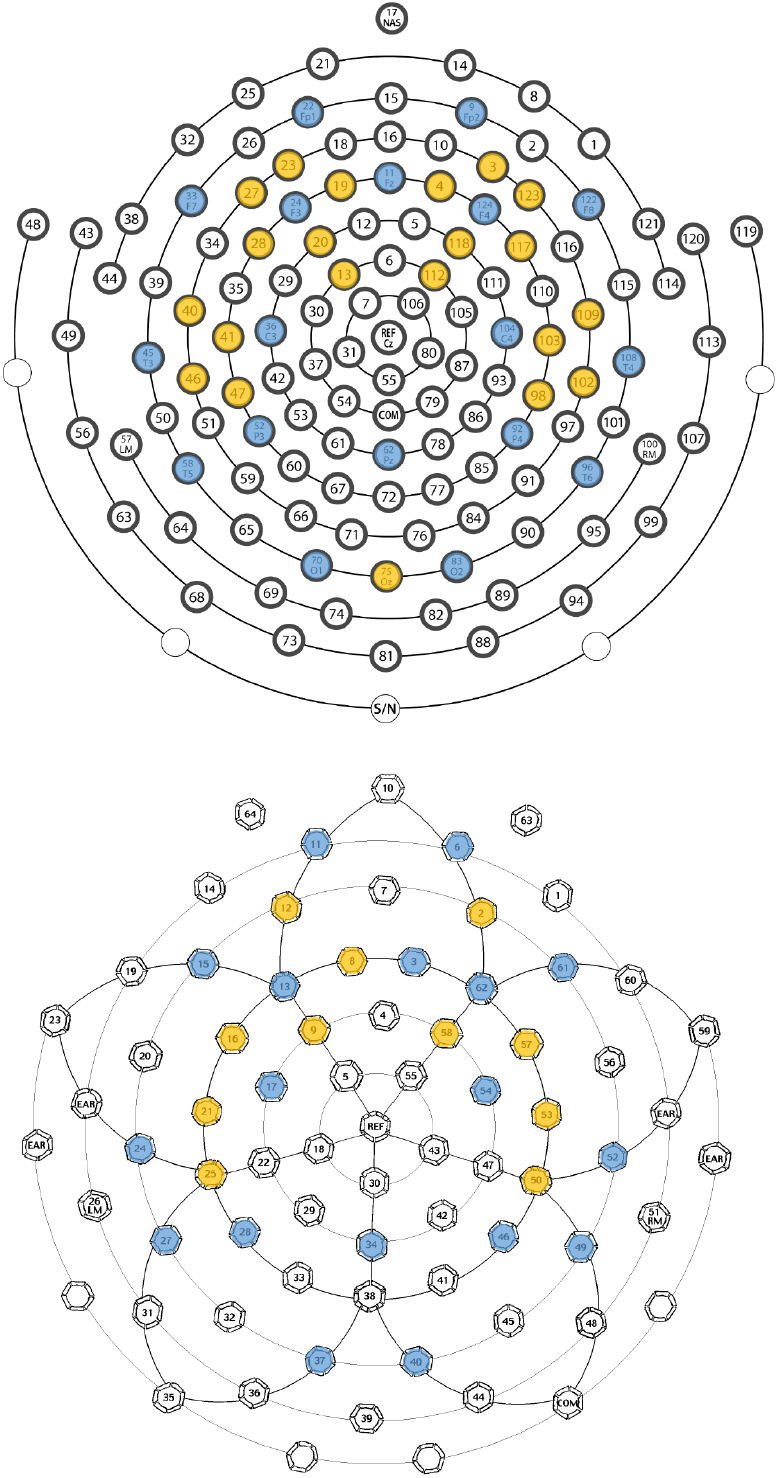
EEG net layouts. Two EEG nets were used in the study: the 128-channel EGI HydroCel Geodesic Sensor Net (version 1.0) presented on the top and the 64-channel EGI Geodesic Sensor Net (version 2.0) presented on the bottom. The 10-20 montage channels evaluated in this study are highlighted in blue, and HAPPE channels included in preprocessing steps are highlighted in yellow.

EEG pre-processing consisted of the following steps for each file. A 1 Hz digital high-pass filter and a 100 Hz low-pass filter were applied to each file, and data sampled at 500 Hz were resampled with interpolation to 250 Hz (so all files had the same effective sampling rate). Bad channels, including channels with high impedances or displacement during recording, were identified using joint probability criteria in HAPPE and were removed from further processing. Artifact removal steps utilizing HAPPE involved removal of 60 Hz electrical noise using CleanLine’s multi-taper approach (Mullen, 2012) and removal of participant artifact rejection such as eye blinks, movement, and muscle activity using wavelet-enhanced ICA with automated component rejection via the Multiple Artifact Rejection Algorithm (Winkler et al., 2014). The bad channels that were removed before artifact rejection were repopulated using spherical interpolation of post-artifact rejection data to reduce spatial bias in rereferencing. EEG data were then rereferenced to the average reference and mean signal detrended.

### EEG data decomposition

After pre-processing with HAPPE, the middle 20 seconds of the longest stretch of consecutive standard phoneme (English ‘da’) presentations in each file were selected for analysis. A 20-second duration was selected in order to maximize the number of language-task participants for inclusion in the study while maintaining length long enough to ensure that the nonlinear time series measures could be appropriately calculated. EEG data from 10-20 channel equivalents (18 channels) for each net type were then decomposed into frequency sub-bands using a discrete wavelet transform and a coarse-graining procedure (described below).

Discrete wavelet transform localizes a signal in both the time and frequency domains, and it is a useful tool that allows analysis of narrow frequency sub-bands of the original signal thought to have biological relevance. The discrete wavelet transform is a filter that decomposes the signal into high pass (details) and low pass (approximations) sub-bands, which are down-sampled to half of the sampling rate. Note that an upper and lower bound of 100 Hz and 1 Hz, respectively, were imposed on the signal in the preprocessing signal steps before wavelet decomposition. The discrete wavelet transform was performed using the PyWavelet *dwt* function with the Daubechies 4 wavelet and symmetric signal extension on the approximate signal of the previous level of decomposition, starting with the original signal at Level 0. Table 5 lists the wavebands that correspond to frequency ranges of wavebands used in traditional EEG power spectral analyses. Only these non-overlapping frequency bands resulting from the wavelet decomposition–D1, D2, D3, D4, D5, and A5–were selected for further analysis with nonlinear measures and covered the range from 1-100 Hz.

**Table 5.** Wavelet decomposition frequency bands for wavelet levels associated with traditional EEG wavebands.

| **Wavelet level** | **Wavelet frequency range** | **Approximate EEG waveband** |
| --- | --- | --- |
| **D1** | 62.5 - 100 Hz | high gamma |
| **D2** | 31.3 - 62.5 Hz | low gamma |
| **D3** | 15.6 - 31.3 Hz | beta |
| **D4** | 7.8 - 15.6 Hz | alpha |
| **D5** | 3.9 - 7.8 Hz | theta |
| **A5** | 1-3.9 Hz | delta |

In addition to the wavelet decomposition, a coarse-grained time series was computed from the original signal using the multiscale entropy analysis methodology introduced by Costa and colleagues *(63)*. The coarse-graining procedure involves dividing the time series into nonoverlapping windows of size *t* and averaging data points within each window, resulting in a signal that is 1/*t* times the length of the original signal. Each scale corresponds to window size: the second scale coarse grains with *t* of two, the third with three, and so on. Five scales were computed from the signal of each electrode, and only multiscale entropy was computed from each of the sub-signals. We note that the coarse-graining procedure is equivalent to using the overlapping frequency bands obtained from the approximations in a discrete wavelet transform using a Haar basis (Bosl, 2017).

### EEG measures

12 different measures were computed for each waveband from each of the 18 10-20 electrodes for each participant. The nonlinear measures quantify the complexity and regularity of the time series using different approaches, and power quantifies the amplitude of oscillations, both on the frequency bands described in Table 5. Table 6 provides a summary description of each measure and credits the software package used for computation.

**TABLE 6:** Descriptions of measures.

| Nonlinear variable | Abbreviation | Description |
| --- | --- | --- |
| **NOLDS software package (64)** |  |  |
| **Detrended fluctuation analysis** | DFA | Measures long-range correlation of the physiological time series |
| **Sample entropy** | SampE | Measures the irregularity of physiological time series without self-matches |
| **Hurst Exponent** | HurstE | Measures the long-term memory processes of a time series |
| **Lyapunov exponent** | LyapE | Measures the chaotic or periodic properties of a time series |
| **EntroPy (https://github.com/raphaelvallat/entropy)** |  |  |
| **Permutation entropy** | PermE | Measures the information content of a given time series based on probability distribution of a set of continuous points |
| **Spectral entropy** | SpecE | Measures degree of skewness in the frequency distribution |
| **Singular value decomposition entropy** | SVDE | Measures the dimensionality of a time series. |
| **Approximate entropy** | AppE | Measures the regularity of times series fluctuations |
| **Higuchi fractal dimension** | HFD | Measures self-similarity in time series using increasingly distanced samples in time |
| **Katz fractal dimension** | KFD | Measures complexity and self-similarity in time series using consecutive time points |
| **Lempel-Ziv Complexity Reference (65)** |  |  |
| **Lempel Ziv Complexity** | LZC | Measures the randomness of finite sequences |
| Linear variable | Abbreviation | Description |
| **SciPy Signal Processing (66)** |  |  |
| **Power** | Power | Measures the frequencies of oscillations in a time series. |

### Feature Selection Strategies

Frequently in clinical modeling, the number of features greatly outnumbers the number of samples in the dataset. In this study we included many different measures in order to provide the most comprehensive description of the EEG signal as possible, resulting in over 1,200 features to describe each participant. However, including an excessive number of features relative to the number of samples leads to model overfitting, which would decrease the model’s generalizability. Therefore, feature selection, or dimensionality reduction, was used in order to retain only the most discriminative features. We evaluated three methods of feature selection using *scikit-learn* software in order to select only the most relevant, informative features: Pearson correlation coefficient, ANOVA F-value, and recursive feature elimination (RFE). The three methods are distinct in that they are based on feature relationship directly with autism outcomes, with variance associated with autism outcomes, and directly with model development, respectively. Each feature selection method was restricted to selecting 20 of the most salient features based on relatedness to class prediction criteria.

### Classification strategies

Multiple classifiers were evaluated in this study, both to identify the best-performing algorithm but also to examine variance across classifier algorithms to see whether they converged on similar prediction performance. There are two main foundational types of supervised learning models, discriminative and generative, that predict the conditional probability of an outcome given input features. Generative models model the distribution of the individual classes with a joint probability distribution. Discriminative models, on the other hand, learn explicit boundaries that best separate the given classes with the conditional probability distribution. We evaluated two generative classifiers, the Naïve Bayes algorithm and linear discriminant analysis, and one discriminative model, an SVM with radial basis functions. Additionally, we included k-nearest neighbors, which is considered a discriminative classifier as well but really employs a voting method based on the outcomes of the *k* nearest neighbors in feature space using a simple Euclidean distance metric. The number of neighbors was set to 7, the square root of the sample size of the 6-month and matching 12-month datasets, and we maintained a selection of 7 neighbors for the full 12-month dataset for consistency. All other models were trained using default parameters from the Python open source package *scikit-learn*. All feature selection methods and machine learning algorithms were used for each dataset analysis, resulting in evaluation of 12 different classification schemes.

### Evaluation of classification strategies

Nested leave-one-out cross validation was used to test the generalizability and validity of model performance given features selected by one of the three dimensionality reduction methods. This validation method involves removing one sample from the dataset, conducting feature selection with the remaining samples to reduce the dimensionality to 20 features, training a model using these samples based on only the selected features, and using that model to predict the outcome of the sample that was removed. This process is repeated for all samples, resulting in one prediction for each sample by a model trained using all other samples. “Nesting” the feature selection method within each iteration of cross validation ensures that the left-out sample in no way contributes to the model that will be used to predict its outcome. Previous studies have found that including test samples in feature selection biases accuracies *(67)*.

There are four possible prediction outcomes: true positive (TP), true negative (TN), false positive (FP), and false negative (FN). Evaluation of model performance in predicting the left-out cases is critical to understanding the ability of a particular algorithm to use learned properties of data to predict the outcome of unseen samples. Five performance criteria (defined below) were calculated for each classifier algorithm:

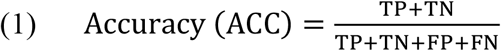

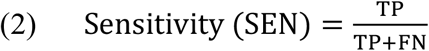

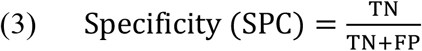

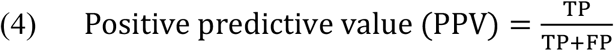

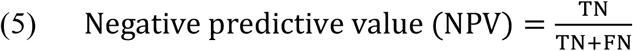

### Characterizing Selected Features Across 6 and 12 months

To determine the average number of times characteristics of a feature (i.e. channel, frequency band, EEG measure) were selected during the nested leave-one-out procedure (Figure 1), the 20 features selected over each of the 54 iterations for 6-month and matching 12-month data sets were collated by the above characteristics, summed, and then divided by 54, the number of iterations/participants. To determine which 20 features were directly compared between outcome groups for each of the above datasets (Table 3), we identified the 20 most frequently selected features over all the iterations. Student’s t-test with Bonferroni correction for multiple comparisons was used to determine if there were significant differences between mean feature values between HR-noASD and HR-ASD groups.

## Data Availability

Python and Jupyter Notebook scripts used for computation of nonlinear measures and machine learning analysis are available. The datasets analyzed during the current study are available from the corresponding author on reasonable request.

## Acknowledgments

We thank all of the families who participated in this study. We also thank Dr. Casey Lew-Williams for his thoughtful feedback and support of this project. Finally, we thank the entire ISP study team for their role in data collection and study coordination.

## Author contributions

C.A.N. and H.T.F. designed the longitudinal study. L.G.-D. and C.L.W. processed the electrophysiological data. F.C.P., C.L.W., L.G.-D., and W.B. designed and carried out the analyses. F.C.P. and W.B. authored the code. F.C.P. drafted the paper, and all authors provided critical revisions.

